# Comparison of DaxibotulinumtoxinA-lanm (Daxi) Versus OnabotulinumtoxinA (BtxA) for Adductor Type Laryngeal Dystonia (AdLD)

**DOI:** 10.1101/2025.05.21.25328112

**Authors:** Alexandra D. D’Oto, Claire E. Perrin, Camryn R. Marshall, VyVy N. Young, Tanvi Rawat, Desi Gutierrez, Sarah L. Schneider, Elizabeth A. Shuman, Clark A. Rosen

## Abstract

**Objectives:** Botulinum toxin A (BtxA) injection for AdLD is typically performed approx. 3 months. DaxibotulinumtoxinA-lamn (Daxi) is a peptide-formulated neuromodulator with reported longer therapeutic duration. This study compared safety and efficacy of Daxi to historical BtxA results in AdLD patients.

**Methods:** In this open-label, prospective clinical trial, 22 stable-dose, BtxA-responsive AdLD patients underwent Daxi injection using manufacturers dosing conversion. To optimize safety, the first 10 patients doses were given in staged fashion (3-6 days between half doses). Safety and duration of voice benefit (DVB) were the primary outcomes. PROMs and voice analyses were obtained pre- and monthly post-injection.

**Results:** Of 22 patients, 20 (75 percent female, mean age= 58.3) were analyzed, with two excluded for injection (misses). No adverse reactions were reported. EAT-10 showed no difference pre- and post-Daxi injection (p=0.068). VHI-10 significantly improved (p=0.004) pre- to post-injection. Procedural LEMG quantitative data for Daxi and BtxA were comparable (p=0.279). DVB of Daxi was longer than BtxA in 8/20 (40 percent), equal to BtxA in 7/20 (35 percent), and shorter than BtxA in 5/19 (25 percent). Those with Daxi benefit had on average 43.9 percent (39.5 days) longer therapeutic duration than their previous BtxA treatments. 8 patients (36.4 percent) returned to BtxA at subsequent injection whereas 13 patients (59.1 percent) desired repeat Daxi injection.

**Conclusion:** This study represents the first use of Daxi for AdLD. Daxi was notably safe and effective, with 40 percent of patients reporting substantive longer duration of voice benefit than with BtxA. Mean duration of voice benefit was 43.9 percent longer than their previous BtxA treatments.

## Introduction

Dystonia is a chronic neurological disorder involving abnormal central motor processing, leading to task-specific muscle spasms triggered by certain actions.^1^ Spasmodic or laryngeal dystonia (LD) is a type of focal dystonia that affects the larynx.^2^ LD is reported to affect around 50,000 individuals in North America, although the incidence may be higher given the high likelihood of misdiagnosis and underdiagnosis in these patients.^3^ Accurate classification of LD is crucial for effective treatment and is generally categorized into three types: adductor-type, abductor-type, and mixed. Adductor-type laryngeal dystonia (AdLD), which constitutes about 80% of cases, is a lifelong condition characterized by disruptive voice breaks due to abnormal adduction of the true vocal folds.^1^ Long-standing theories suggest that AdLD may be related to a sensory processing abnormality, which was recently supported by findings published by Young et al. demonstrating increased laryngeal sensation in AdLD patients compared to controls. ^4^ Management strategies aim to alleviate symptoms through medical or procedural intervention. The gold standard treatment for AdLD is the injection of botulinum toxin (BtxA) into the affected muscles, specifically the thyroarytenoid-lateral cricoarytenoid (TA-LCA) complex.^5^ BtxA, available in products such as Botox® and Dysport®, has been shown to improve voice quality by 70-80%, with benefits lasting three to four months.^5^

Botulinum toxin type A (BtxA) is a naturally occurring neurotoxin that temporarily inhibits the pre-synaptic release of acetylcholine at neuromuscular junctions, resulting in reversible muscle paralysis. Following injection, patients may experience an initial period of breathiness due to TA-LCA complex weakness prior to symptomatic relief. Some studies suggest that a greater degree of initial TA-LCA complex weakness may correlate with a longer duration of benefit, although this must be balanced with the overall functionality of the patient’s voice.^6^ Therapeutic benefit from BtxA injections typically lasts for 3-4 months, with breathiness lasting from a few days to a few weeks shortly after injection.

DaxibotulinumtoxinA (Daxi) is a recent formulation of botulinum A toxin utilizing a proprietary, stabilizing peptide (RTP004) and polysorbate-20, as opposed to human serum albumin (HSA) as a stabilizer in standard formulations of BtxA.^7^ The positive charge of the peptide complex to the botulinum toxin is thought to promote binding at the negatively charged presynaptic terminal, allowing for less diffusion of the neurotoxin complex.^8,9^ Further, this enhanced binding has been shown in studies to prolong anchoring at the presynaptic terminal.^9^ Botulinum toxin A bound to the peptide complex in Daxi is thus theorized to remain present at the presynaptic terminal longer than the standard HSA formulation of BtxA, resulting in a potentially longer duration of benefit.^7,9^ In May 2023, the United States Food and Drug Administration (FDA) approved Daxi for the treatment of moderate to severe glabellar lines and cervical dystonia. In clinical trials, Daxi demonstrated a duration of benefit lasting 6 to 9 months in the treatment of glabellar lines.^10^ Additionally, Daxi was studied in clinical trials for the treatment of moderate to severe idiopathic cervical dystonia.^11^ The median duration of effect, or time to at least 80% loss of peak improvement, was 24 weeks with Daxi 150U and 20.3 with Daxi 250U, with no significant difference between these doses.^11^ The aforementioned clinical trials report a benefit period significantly longer than the documented 12-16 weeks of benefit with BtxA reported by the manufacturer.^12^ Notably, Daxi demonstrated a safety profile similar to BtxA in all studies.^10,11,13^ Daxi also has been used off-label in lieu of BtxA for several other medical conditions, including chronic migraine, spasticity, and respiratory dystonia, to investigate Daxi’s utility in providing therapeutic benefit while extending treatment intervals.^8,14^

To date, there are no studies exploring the use of Daxi in AdLD patients. This study represents the first investigation to compare the safety and efficacy of Daxi compared to BtxA in AdLD treatment.

## Materials and Methods

After obtaining Institutional Review Board approval, an open-label, prospective clinical trial (NCT05158166) was developed at the University of California San Francisco Voice and Swallowing Center. Stable-dose, BtxA-responsive patients > 18 years of age with a diagnosis of AdLD were included in the study. Stable-dose patients were defined as those who had received and benefitted from the <u>same</u> BtxA dosing for their past 3 BtxA injections. Patients were excluded from the study if they were diagnosed with other neurologic conditions, including abductor-type laryngeal dystonia, amyotrophic lateral sclerosis, multiple sclerosis, Parkinson’s disease, and essential tremor. Informed consent was obtained from enrolled patients for the administration of DaxibotulinumtoxinA-lanm (DAXXIFY®, Revance Therapeutics, Inc., Nashville, TN) in lieu of BtxA.

Daxi injections were performed by a fellowship-trained laryngologist with ≥14 years’ experience utilizing quantitative laryngeal electromyography (LEMG) to aid in TA-LCA complex identification.^15^ The manufacturer’s recommended dosing conversion from BtxA to Daxi was used after personal communication with Revance Therapeutics, Inc (July 2024). Prior clinical trials also used a 2:1 BtxA to Daxi dose conversion.^10,11^ Through this dosing conversion, patients’ BtxA doses were doubled to determine Daxi doses. To establish safety and monitor for adverse effects, the first 10 patients’ planned doses were divided into half doses and administered 3-6 days apart with direct follow-up regarding voice and breathing symptoms after the first half dose prior to proceeding with the second half dose.

Safety and duration of voice benefit (DVB) were the primary outcomes. DVB was determined by calculating the difference of time (in days) between injection date and time the patient reported a significant reduction of benefit that would prompt scheduling a repeat treatment. DVB is routinely calculated for all AdLD patients during their return visits by asking “how many days ago did you lose injection benefit such that you felt you needed another injection.” This time frame was then subtracted from number of days between injection dates to calculate DVB. For BtxA, a mean DVB was calculated by averaging the last 3 DVB values to compare to the Daxi DVB. Clinically significant difference in DVB was *a priori* determined as >14 days between values to allow for normal inter-injection variability.

Patient reported outcome measures (PROMs) and voice analyses were obtained pre-Daxi injection and monthly post-Daxi injection. REDCap (LTS Version 14.5.32, Vanderbilt University) was used for patient data entry of Voice Handicap Index–10 (VHI-10)^16^, Eating Assessment Tool (EAT-10)^17^, Communicative Participation Item Bank-10 (CPIB-10)^18^, and OMNI-Voice Effort Scale (OMNI-VES)^19^ questionnaires. Pre-Daxi injection EAT-10 values were compared to EAT-10 values 5 days post-Daxi injection. Pre-injection VHI-10, CPIB-10, and OMNI-VES data were compared to values collected 30 days post-injection. Phonalyze (Praat Version 6.3, Cognizn LLC, Walnut Creek, CA) was utilized for secure collection and analysis of voice samples via the patients’ smartphones. Two voice-specialized speech-language pathologists with 3-8 years of voice experience were selected to blindly rate standard voice samples using the Consensus Auditory-Perceptual Evaluation of Voice (CAPE-V) for patients pre- and 2 months post-Daxi injections.^20^ Additional perceptual characteristics including voiced (adductor) and voiceless (abductor) breaks were rated along with the standard characteristics, as is advised per the CAPE- V protocol. Smoothed Cepstral Peak Prominence (CPPS) data was also compared between pre- and 2-month post-Daxi injection. LEMG data including the number of small segments (NSS) and turns was recorded for each patient’s Daxi injection and compared to the average of the last 3 BtxA injections.^15^ Prior BtxA injection clinical documentation pertaining to breathiness period, DVB, and dysphagia concerns, along with LEMG data, were retrieved through electronic medical record chart review.

Analyses of voice improvement data comparing BtxA and Daxi injections were performed using Microsoft Excel (Version 16, Microsoft Corporation, Redmond WA). Statistical analyses were performed via SPSS (IBM SPSS Statistics for Macintosh, Version 28.0). An independent t-test was utilized to determine significant differences in objective measures and PROMs between Daxi injections and prior BtxA injections. Inter- and intra-rater reliability was calculated for CAPE-V ratings using Intraclass Correlation Coefficient (ICC) reliability index. ICC value ranges were as follows: 0.00-0.20 (poor agreement), 0.21-0.41 (fair agreement), 0.41-0.60 (moderate agreement), 0.61-0.80 (good agreement), and 0.81-1.00 (excellent reliability).^21^

## Results

Twenty of the 22 enrolled patients (75% female, mean age= 58.3 years) were included in the analysis (Table 1). Two patients had no change in their voice and no swallowing impact. These patients were thus considered “misses,” or injection events that did not reach the intended TA-LCA complexes and thus were not included in the subsequent analysis. No adverse events were reported in the entire study cohort. No significant difference was noted for either NSS or turns on LEMG (p=0.279), denoting consistency between injections. VHI-10 and OMNI-VES scores showed a significant difference between pre- and post-Daxi injections for all patients (p<0.001) (Table 2). There was no significant difference in pre- and post-Daxi injection CPIB (p=0.153) or EAT-10 (p=0.068).

**Table 1:** Patient Demographic Information for Daxi Cohort.

| **Patient** | **Age Range** | **Gender** | **Ethnicity** |
| --- | --- | --- | --- |
| 1 | 45-50 | Male | White |
| 2 | 65-70 | Male | White |
| 3 | 70-75 | Female | White |
| 4 | 55-60 | Female | White |
| 5 | 30-35 | Female | White |
| 6 | 55-60 | Male | White |
| 7 | 55-60 | Female | White |
| 8 | 60-65 | Female | White |
| 9 | 55-60 | Male | White |
| 10 | 70-75 | Female | White |
| 11 | 40-45 | Female | Native Hawaiian/Alaskan |
| 12 | 65-70 | Female | White |
| 13 | 25-30 | Female | White |
| 14 | 70-75 | Female | White |
| 15 | 70-75 | Female | White |
| 16 | 50-55 | Female | White |
| 17 | 50-55 | Female | White |
| 18 | 70-75 | Female | Black/African American |
| 19 | 40-45 | Female | White |
| 20 | 70-75 | Female | White |

\*Daxi: DaxibotulinumtoxinA-lanm

**Table 2:** Patient Reported Outcome Measures at Baseline and 30 Days Post-Daxi Injection.

| **Outcome** |  |  | **p-value** |
| --- | --- | --- | --- |
| VHI-10 | Baseline | 21.10±6.82 | p<0.001 |
|  | 30 days | 17.81±8.18 |  |
| EAT-10 | Baseline | 4.60±5.66 | p=0.068 |
|  | 30 days | 5.16±5.94 |  |
| CPIB | Baseline | 15.15±6.27 | p=0.153 |
|  | 30 days | 16.69±7.08 |  |
| OMNI-VES | Baseline | 5.95±2.01 | p<0.001 |
|  | 30 days | 3.69±2.39 |  |

\*Daxi: DaxibotulinumtoxinA-lanm
VHI-10: Voice Handicap Index 1-
EAT-10: Eating Assessment Tool
CPIB: Communicative Participation Item Bank
OMNI-VES: OMNI-Vocal Effort Scale

Six patients did not complete the 2-month post-injection CAPE-V. Thus, 14 subjects total were available for complete CAPE-V analysis pre- and post-Daxi (Table 3). On analysis of recordings, Roughness improved from baseline at 2 months post-Daxi injection (p=0.026), with mean scores between the raters decreasing from 39.8 to 29.1. Improvement in Voiced Breaks approached but did not reach significance (p=0.053), with mean score reduction from 29.0 to 25.3. There was no significant change in Overall Severity (p=0.183), Breathiness (p=0.09), or Strain (p=0.475). Intra-rater reliability was 0.879 for Rater 1 and 0.94 for Rater 2 (both excellent reliability). The ICC value for overall severity rating between the raters was 0.89 (excellent reliability).

**Table 3:**
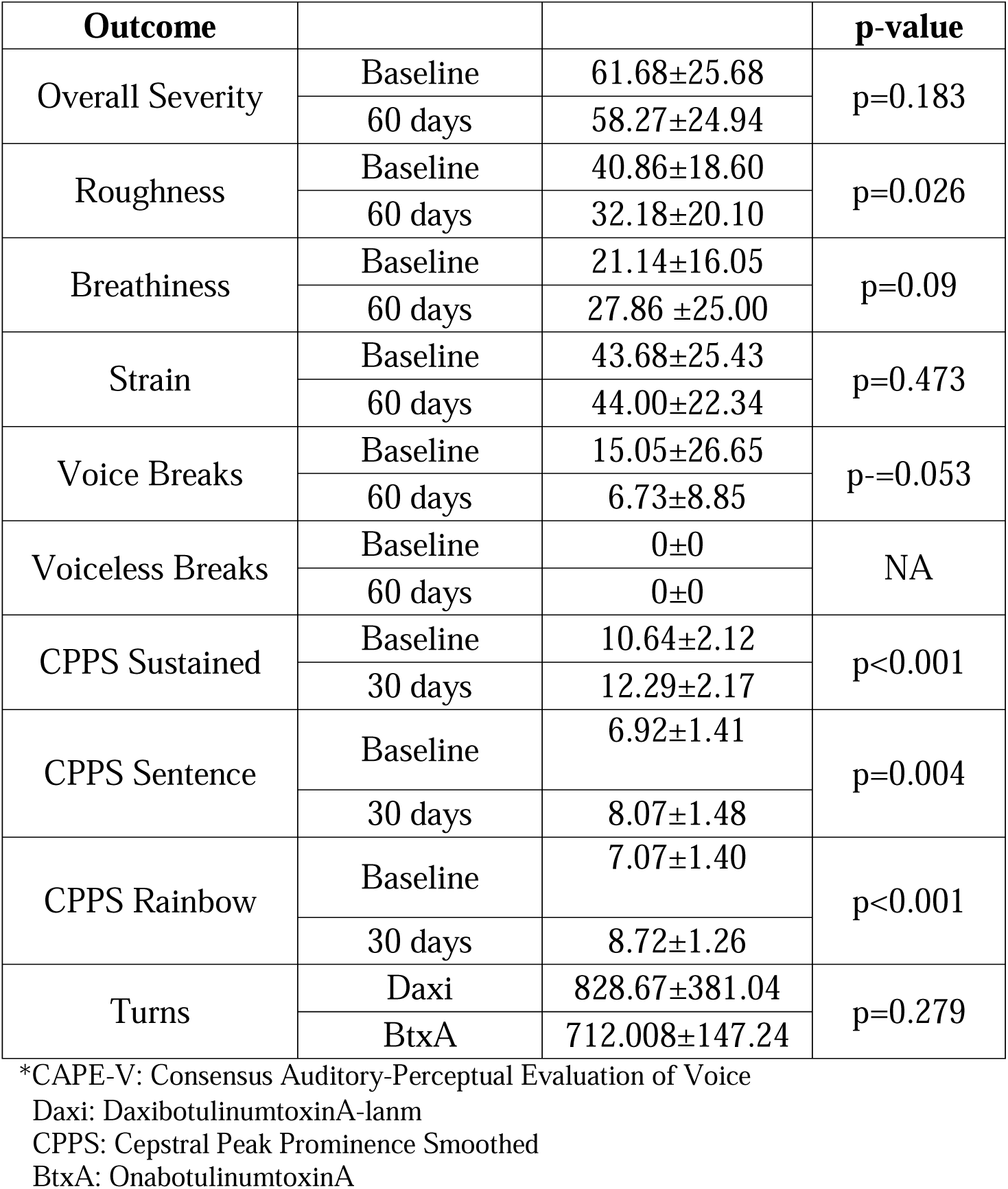
Objective and CAPE-V Outcomes at Baseline and 60 Days Post-Daxi Injection.

CPPS was obtained for sustained vowel phonation, CAPE-V voiced sentence (“we were away a year ago”) reading, and rainbow passage reading at baseline (BL) and 2-month post-Daxi injection (PI) (Table 3). Seven patients did not complete the PI sustained vowel phonation or voiced sentence recordings, and 10 patients did not complete the PI rainbow passage reading. Fourteen subject recordings underwent CPPS analysis for the sustained vowel phonation and voiced sentence, while ten underwent CPPS analysis for rainbow passage reading. There was significant improvement in post-injection sustained vowel phonation (p<0.001, BL range = 4.91-14.59; PI range = 10.00-15.01), CAPE-V sentences (p=0.004, BL range = 4.71-9.16; PI range = 5.8-11.24), and rainbow passage reading (p=<0.001, BL range = 4.79-9.04; PI range = 6.44-10.72) (Table 3).

Duration of voice benefit was found to be variable within the study cohort (Figure 1, Table 4). In 8 of 20 patients (40%), DVB for Daxi was longer than DVB for BtxA, demonstrating average 130.6 days (range= 34-179 days) with Daxi compared to 86.1 days (range= 51-111 days) with BtxA (p<0.001). Those who experienced a longer DVB with Daxi (Group A) had on average 43.9% (39.5 days) longer therapeutic duration than with their previous BtxA injections. Interestingly, 7 patients (35%) had equivalent DVB with Daxi (Group B) (mean= 86.86 days, range= 69-118 days) compared to their prior BtxA injections (mean= 83.29 days, range= 70-111 days, p=0.04). Finally, compared to prior BtxA treatment (mean= 81.8 days, range= 52-92 days), five patients (25%) reported shorter DVB with Daxi (Group C) (mean= 49.8 days, range= 37-66 days, p=0.001). There was no significant difference in post-injection breathiness period between Daxi (average= 9.55 days, range= 0-30 days) and BtxA (average = 11.57 days, range= 0-30 days, p=0.169).

**Figure 1:**
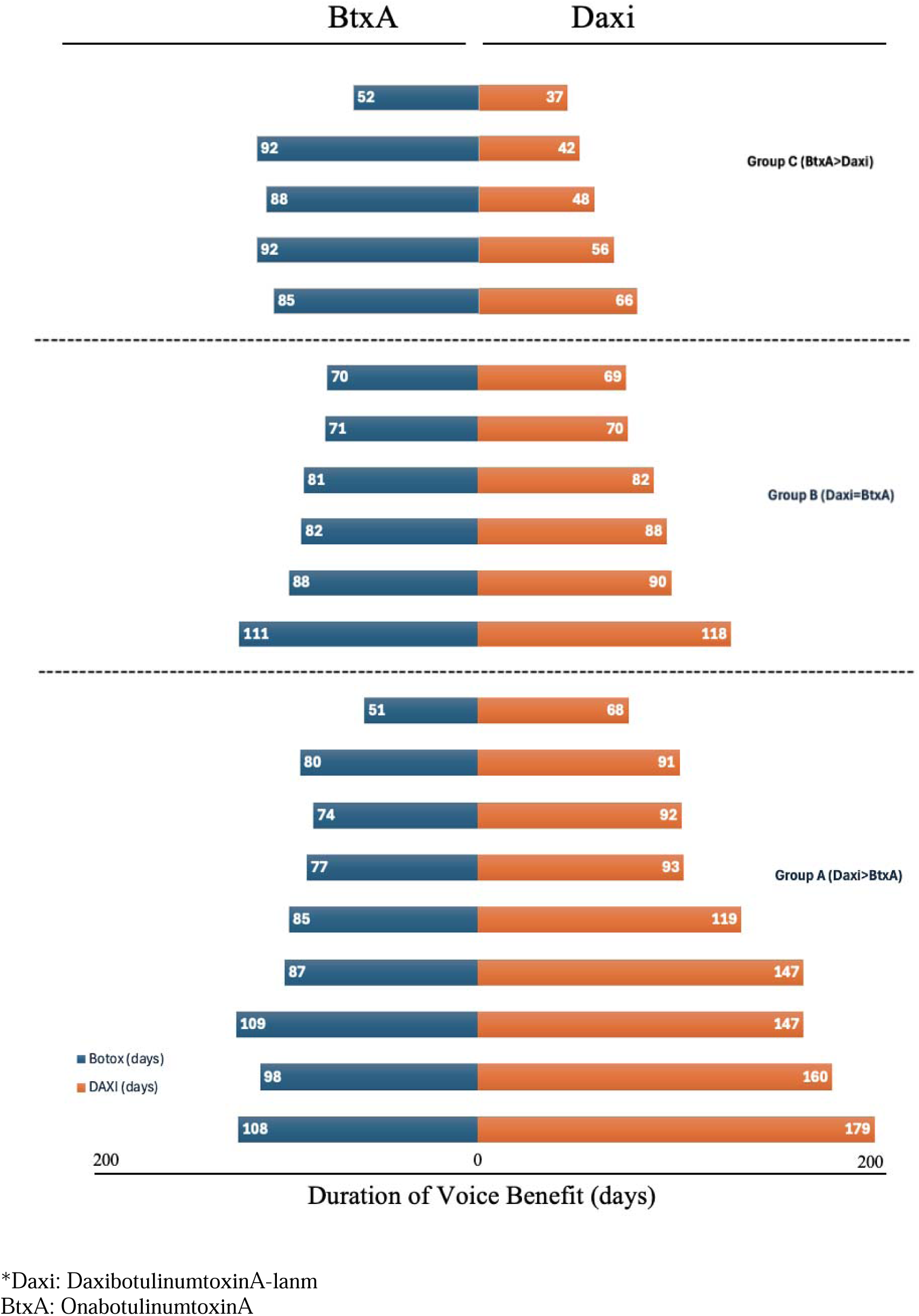
Comparison of Duration of Voice Benefit in Days Between Daxi and BtxA.

**Table 4:**
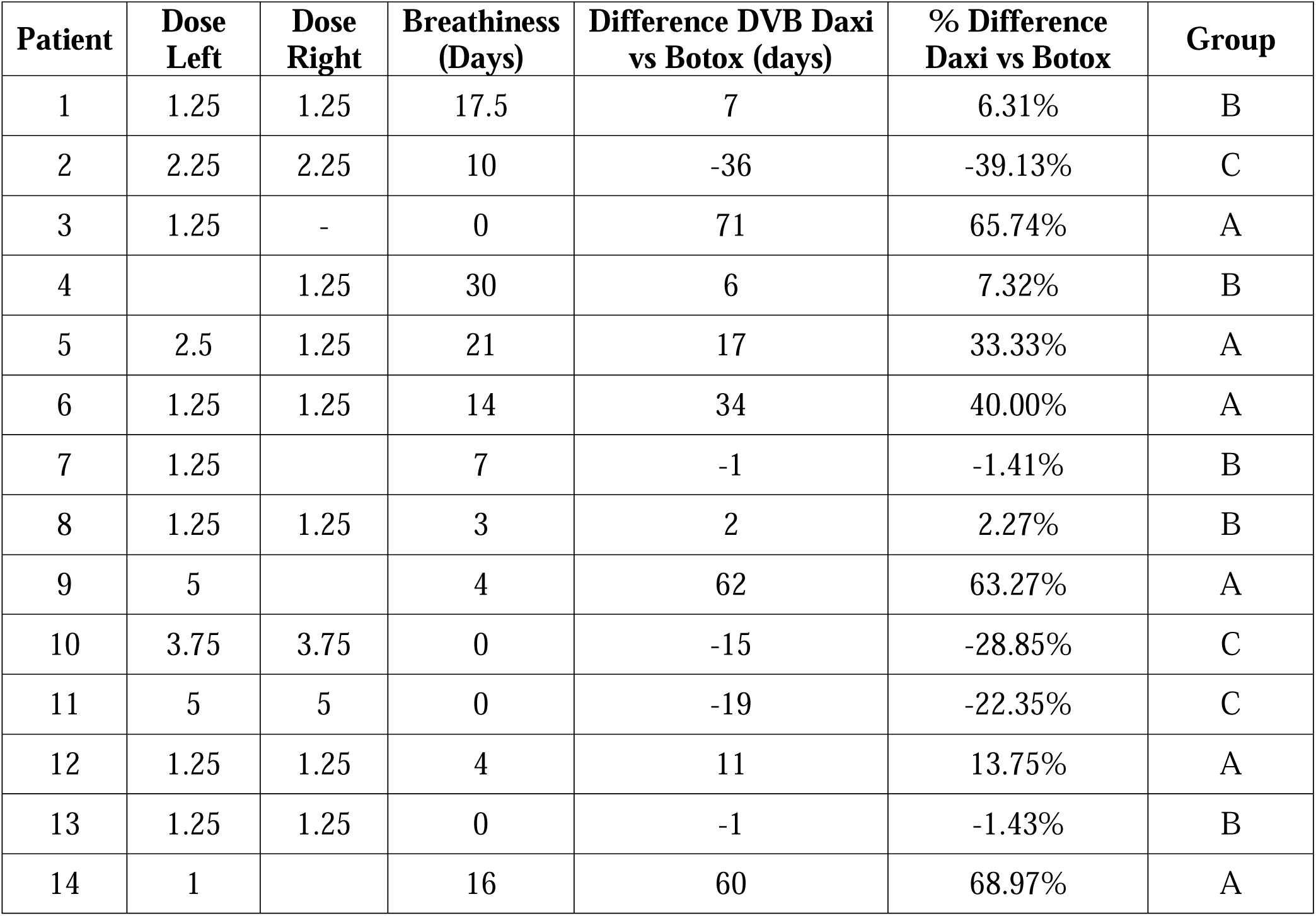
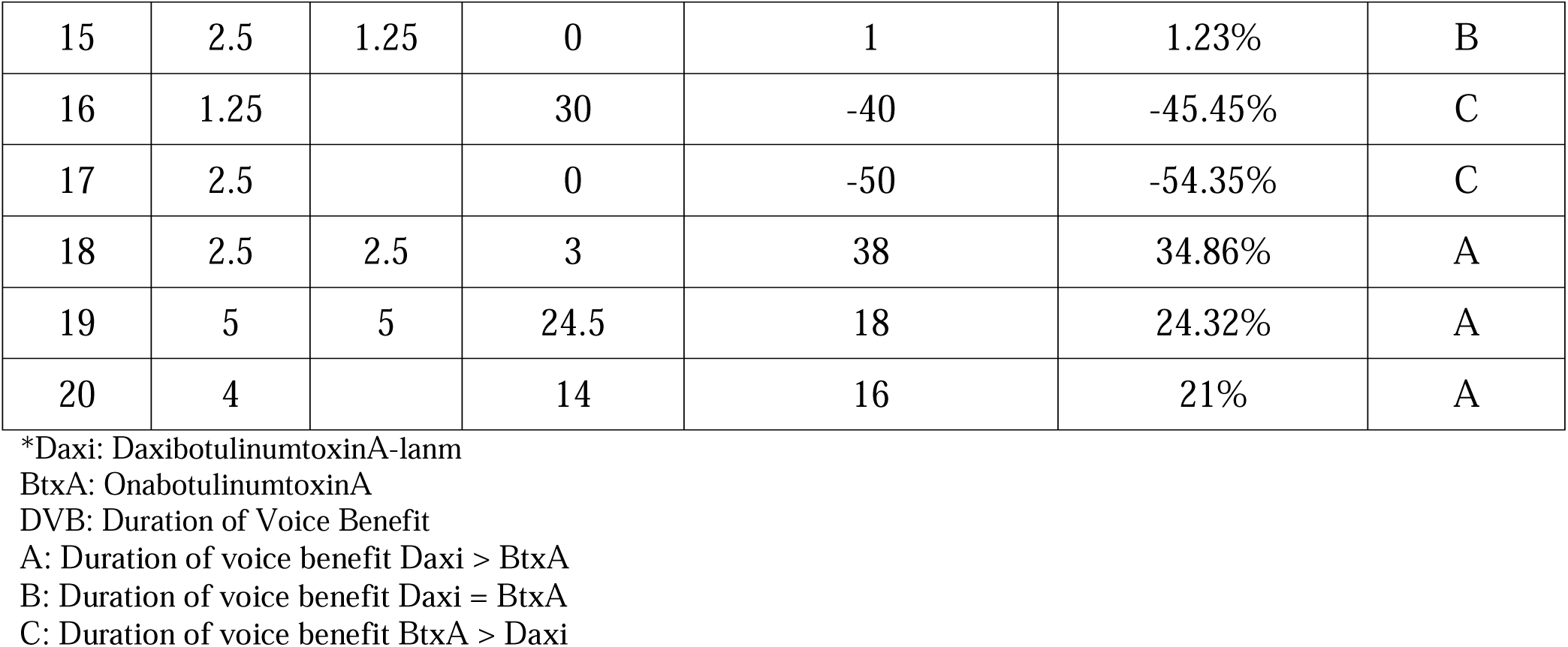
Daxi Dosing and Comparison of Breathiness Duration, and Voice Benefit Differences Relative to BtxA.

| **Patient** | **Dose Left** | **Dose Right** | **Breathiness (Days)** | **Difference DVB Daxi vs Botox (days)** | **% Difference Daxi vs Botox** | **Group** |
| --- | --- | --- | --- | --- | --- | --- |
| 1 | 1.25 | 1.25 | 17.5 | 7 | 6.31% | B |
| 2 | 2.25 | 2.25 | 10 | -36 | -39.13% | C |
| 3 | 1.25 | - | 0 | 71 | 65.74% | A |
| 4 |  | 1.25 | 30 | 6 | 7.32% | B |
| 5 | 2.5 | 1.25 | 21 | 17 | 33.33% | A |
| 6 | 1.25 | 1.25 | 14 | 34 | 40.00% | A |
| 7 | 1.25 |  | 7 | -1 | -1.41% | B |
| 8 | 1.25 | 1.25 | 3 | 2 | 2.27% | B |
| 9 | 5 |  | 4 | 62 | 63.27% | A |
| 10 | 3.75 | 3.75 | 0 | -15 | -28.85% | C |
| 11 | 5 | 5 | 0 | -19 | -22.35% | C |
| 12 | 1.25 | 1.25 | 4 | 11 | 13.75% | A |
| 13 | 1.25 | 1.25 | 0 | -1 | -1.43% | B |
| 14 | 1 |  | 16 | 60 | 68.97% | A |
| 15 | 2.5 | 1.25 | 0 | 1 | 1.23% | B |
| 16 | 1.25 |  | 30 | -40 | -45.45% | C |
| 17 | 2.5 |  | 0 | -50 | -54.35% | C |
| 18 | 2.5 | 2.5 | 3 | 38 | 34.86% | A |
| 19 | 5 | 5 | 24.5 | 18 | 24.32% | A |
| 20 | 4 |  | 14 | 16 | 21% | A |

\*Daxi: DaxibotulinumtoxinA-lanm
BtxA: OnabotulinumtoxinA
DVB: Duration of Voice Benefit
A: Duration of voice benefit Daxi > BtxA
B: Duration of voice benefit Daxi = BtxA
C: Duration of voice benefit BtxA > Daxi

Thirteen of twenty-two patients (59.1%) desired repeat Daxi injection at their subsequent visit, while eight patients (36.4%) returned to their previous BtxA dosing. Two of these 8 patients had equivocal Daxi DVB compared to BtxA DVB, while 3 had worse Daxi DVB compared to BtxA DVB. The remaining 2 patients were the aforementioned “misses,” both of whom returned to BtxA injections. One patient (4.5%), who did report significantly longer DVB with Daxi, was unable to participate in repeat Daxi injection as the subsequent injection fell outside of the clinical study window. The patients who were eligible and opted for additional Daxi injections were offered to participate in a second related clinical trial (dose consistency) and/or a third related clinical trial (dose escalation). These results and findings will be reported in a future publication.

## Discussion

AdLD is a neurological voice disorder with considerable impact on patient quality of life.^22^ The ability to meet occupational and social vocal demands relies heavily on clear and effective communication. Those with AdLD can be significantly hindered by their dysphonia in their day-to-day interactions. As a result, AdLD patients may have negative social and occupational encounters due to their voice pattern.^23^ Those affected may be perceived as nervous or anxious, as well as difficult to understand due to frequent involuntary vocal breaks.^23^ In Isetti et al., AdLD was found to contribute to presenteeism, or decreased ability to perform work due to a health condition.^24^ Moreover, patients expressed concerns regarding job demotion and position restructuring due to their AdLD.^24^ The communication barrier can thus lead to difficulty in securing and maintaining employment.^23^

The current mainstay of treatment for AdLD is chemodenervation of the TA-LCA complex with BtxA. Although BtxA is efficacious, its effect is transitory, resulting in retreatment every 3-4 months. The frequency of repeated injections can contribute to AdLD patients’ treatment burden. Young and Halstead demonstrated that AdLD patients with lower socioeconomic status received fewer BtxA injections thought to be due in part to increased out-of-pocket costs despite insurance status.^25^ Additionally, Hur et al. noted that lower income and racial minority AdLD patients were more likely to forgo care due to lack of transportation.^26^ Treatment access is often limited to academic centers given the rarity of AdLD and need for laryngology expertise. As a result, patients may need to travel long distances to receive BtxA injections. These factors can be detrimental to overall patient well-being, and unsurprisingly, an association between LD and psychological conditions, such as depression and anxiety, has been established in multiple studies. ^23,27,28^

Future treatment for AdLD should aim to reduce the financial and time-consuming aspects of repeated therapy, while maintaining efficacy and reliability. Daxi is a novel preparation of botulinum toxin A that has demonstrated a prolonged duration of benefit in glabellar line and cervical dystonia treatment.^10,11,29^ Clinical trials of Daxi for glabellar lines demonstrated a median duration of therapeutic benefit of 24 weeks with return to baseline at 27.7 weeks.^10^ Similarly, cervical dystonia patients demonstrated lasting treatment response up to 24 weeks compared to their prior 12-16 weeks of benefit.^11,12^ All of these trials, notably, demonstrated a similar side effect profile to BtxA.^10,11,13^

The current study sought to determine the safety and efficacy of Daxi compared to BtxA in the treatment of AdLD. No adverse events were reported, and no significant dysphagia symptoms were encountered as demonstrated by pre- and post-injection EAT-10 results. These findings showcase a similar safety and side effect profile as BtxA, which allows for ease of counseling when considering Daxi vs BtxA injections in AdLD patients. During phase 1 of this prospective clinical trial, Daxi was found to have a significantly longer therapeutic duration of benefit in 40% of patients. Importantly, no increased breathiness period was observed with Daxi injections compared to BtxA injections. Among these patients, the voice benefit lasted on average 43.9% longer than their prior BtxA injections. A 39.5-day prolongation of voice benefit could result in 1 less clinic visit per year for patients who received superior benefit to BtxA with Daxi. Reduction in clinic visits could alleviate the financial and logistical challenges associated with the need for repeated lifetime injections, especially in patients traveling from a great distance to obtain care.

Two of 22 injections (9%) were considered “misses” despite LEMG guidance. There are few studies exploring ineffective injection rates. Fulmer et al. reported a 6.2% “miss” rate with LEMG guidance and 6.5% with point-touch technique.^30^ Barrow et al. noted a 3.36% “miss” rate out of 115 BtxA injections.^31^ Galardi et al., however, noted 30.2% of BtxA injections were ineffective despite LEMG use.^32^ Although the percentage of “misses” in this study falls within the above reported ranges, it is unclear if Daxi would demonstrate a significantly different ineffective injection rate than BtxA. As previously mentioned, Daxi exhibited less diffusion than BtxA at target sites.^9^ While less diffusion is theoretically more beneficial to reduce off-target effects, this may in turn require more precise placement of the neurotoxin to elicit the desired response. Additionally, wide range of Daxi DVB (34-179 days in this study) raises further questions regarding factors that may affect its therapeutic duration. Investigation into patient factors, severity of AdLD, dose titration, consistency of response, and Daxi diffusion rate may shed light on optimization and patient selection for Daxi use in this population.

The significant improvement in VHI-10 and OMNI-VES post-injection demonstrates the efficacy of Daxi in treatment of AdLD and its associated voice symptoms. The significant improvement in post-injection CPPS for sustained vowel phonation, CAPE-V voiced sentence reading, and rainbow passage reading from baseline CPPS objectively supports Daxi’s therapeutic benefit. Although ratings of Roughness improved on the CAPE-V, no significant difference was noted in Overall Severity, Breathiness, or Strain. The incomplete post-injection CAPE-V data likely exacerbates the discrepancy between PROMs and CAPE-V ratings. Seven of 20 patients (35%) did not complete their 2-month recordings for analysis. The lower response rate could be addressed in subsequent studies by reducing PROM-completion burden.

There were several limitations to this study. As this was the first prospective trial investigating Daxi for AdLD, the number of patients enrolled was intentionally kept relatively small. This allowed for more frequent patient check-ins and close monitoring for adverse events. While the smaller cohort allowed for improved safety measures, a larger sample size could potentially reveal rare adverse effects, if any. As no adverse events were reported in the current study, future research with repeated injections, dose escalation, and increased patient enrollment will aid in elucidating the Daxi safety profile as compared to the BtxA safety profile. Another limitation is the Daxi dose was determined based on the manufacturer’s recommendation, which was double the BtxA dose. The dose conversion was based on previous clinical trials comparing the efficacies of Daxi and BtxA in non-laryngeal conditions, such as cervical dystonia and glabellar lines. The proposed dosing conversion did allow for demonstrated clinical benefit using Daxi. However, it is unclear at this time if additional benefit would be gained through further titration and dose escalation. Later phases of the current study will explore the effect of dosing on Daxi treatment response.

## Conclusion

This study represents the first reported use of Daxi for treatment of AdLD. Daxi was found to be safe and effective, with 40% of patients reporting longer duration of voice benefit with Daxi than with their prior BtxA injections by a minimum of 14 days. In those with longer DVB, patients reported a 43.9% longer therapeutic duration with Daxi than experienced with BtxA. Additional phases of the current study will explore the safety and efficacy of Daxi with repeated dose administration and dose escalation in AdLD patients.

## Data Availability

All data produced in the present work are contained in the manuscript.

## Acknowledgements

Revance Therapeutics, Inc., provided free DAXXIFY® and partial financial support as an Investigator Sponsored Trial; however, Revance had no input on study design, study performance or study result analysis.

Study financial support also was partially provided by Dysphonia International, but Dysphonia International had no impact on study design, study performance or analysis of results.

The authors would like to acknowledge Jack Goldstein, PhD, and Sky T. Yang, CCC-SLP, for their contributions to this study.

